# Epidemiology and outcomes of SARS-CoV-2 infection associated with anti-nucleocapsid seropositivity in Cape Town, South Africa

**DOI:** 10.1101/2022.12.01.22282927

**Authors:** Hannah Hussey, Helena Vreede, Mary-Ann Davies, Alexa Heekes, Emma Kalk, Diana Hardie, Gert van Zyl, Michelle Naidoo, Erna Morden, Jamy-Lee Bam, Nesbert Zinyakatira, Chad M Centner, Jean Maritz, Jessica Opie, Zivanai Chapanduka, Hassan Mahomed, Mariette Smith, Annibale Cois, David Pienaar, Andrew D. Redd, Wolfgang Preiser, Robert Wilkinson, Kamy Chetty, Andrew Boulle, Nei-yuan Hsiao

## Abstract

**Background:** In low- and middle-income countries where SARS-CoV-2 testing is limited, seroprevalence studies can characterise the scale and determinants of the pandemic, as well as elucidate protection conferred by prior exposure.

**Methods:** We conducted repeated cross-sectional serosurveys (July 2020 – November 2021) using residual plasma from routine convenient blood samples from patients with non-COVID-19 conditions from Cape Town, South Africa. SARS-CoV-2 anti-nucleocapsid antibodies and linked clinical information were used to investigate: (1) seroprevalence over time and risk factors associated with seropositivity, (2) ecological comparison of seroprevalence between subdistricts, (3) case ascertainment rates, and (4) the relative protection against COVID-19 associated with seropositivity and vaccination statuses, to estimate variant disease severity.

**Findings:** Among the subset sampled, seroprevalence of SARS-CoV-2 in Cape Town increased from 39.2% in July 2020 to 67.8% in November 2021. Poorer communities had both higher seroprevalence and COVID-19 mortality. Only 10% of seropositive individuals had a recorded positive SARS-CoV-2 test. Antibody positivity before the start of the Omicron BA.1 wave (28 November 2021) was strongly protective for severe disease (adjusted odds ratio [aOR] 0.15; 95%CI 0.05-0.46), with additional benefit in those who were also vaccinated (aOR 0.07, 95%CI 0.01-0.35).

**Interpretation:** The high population seroprevalence in Cape Town was attained at the cost of substantial COVID-19 mortality. At the individual level, seropositivity was highly protective against subsequent infections and severe COVID-19.

**Funding:** Wellcome Trust, National Health Laboratory Service, the Division of Intramural Research, NIAID, NIH (ADR) and Western Cape Government Health.

**Research in context:** *Evidence before this study:* In low- and middle-income countries where SARS-CoV-2 testing is limited, seroprevalence studies can help describe the true extent of the pandemic. Infection from Omicron was associated with less severe disease in South Africa, but it is unclear whether this was due to a decrease in virulence of the variant or if prior infection provided protection.

*Added value of this study:* The seroprevalence data nested within a population cohort enabled us to assess differential case ascertainment rates, as well as to examine the contribution of both natural and vaccine-induced immunity in protecting communities against infections and severe disease with different SARS-CoV-2 variants.

*Implications of the available evidence:* Inequality and differential access to resources resulted in poorer communities having higher seroprevalence and COVID-19 death rates, with lower case ascertainment rates. Antibody positivity provided strong protection against an immune escape variant like Omicron but came at a high mortality cost.

## Introduction

Virtually every country in the world, including most low- and middle-income countries experienced multiple waves of COVID-19 cases in the first two years of the pandemic but due to a relative lack of access to testing, the reported cases are a gross under-ascertainment of the true scale of infection. In South Africa, more than 100,000 COVID-19-related deaths have been reported with excess death estimated to be more than 300,000 between March 2020 and April 2022 (1,2).

Recent serosurvey studies in South Africa (3–6) and Africa (7,8) have demonstrated high levels of sero-positivity from infection or vaccination two years after the emergence of SARS-CoV-2. However, due to their cross-sectional design, few large seroprevalence studies have been able to estimate the degree of protection conferred by seropositivity beyond ecological comparisons (3).

A serosurvey that is nested within a population cohort can provide insight into the relative protection of prior infection and vaccination. This is particularly useful in the South African complex immune landscape where many individuals have hybrid immunity from both prior infection with different variants as well as vaccination (10). In this study we use serology with anonymously linked infection and outcome data from a population-level medical database to describe in detail the longitudinal epidemiology of SARS-CoV-2 in Cape Town, South Africa, across three SARS-CoV-2 waves between March 2020 and March 2022. We aimed to assess the protection against future infection and severe disease conferred by different combinations of prior infection and vaccination; and assess differences in the risk for severe disease associated with viral variants in non-immune, unvaccinated individuals prior to each wave, according locally developed wave definition (11). These waves peaked in June 2020, and January, August and December 2021, and were caused by the ancestral SARS-CoV-2, the Beta, Delta and Omicron (BA.1/BA.2 sub-lineages) variants, respectively (Figure 1) (12).

**Figure 1:**
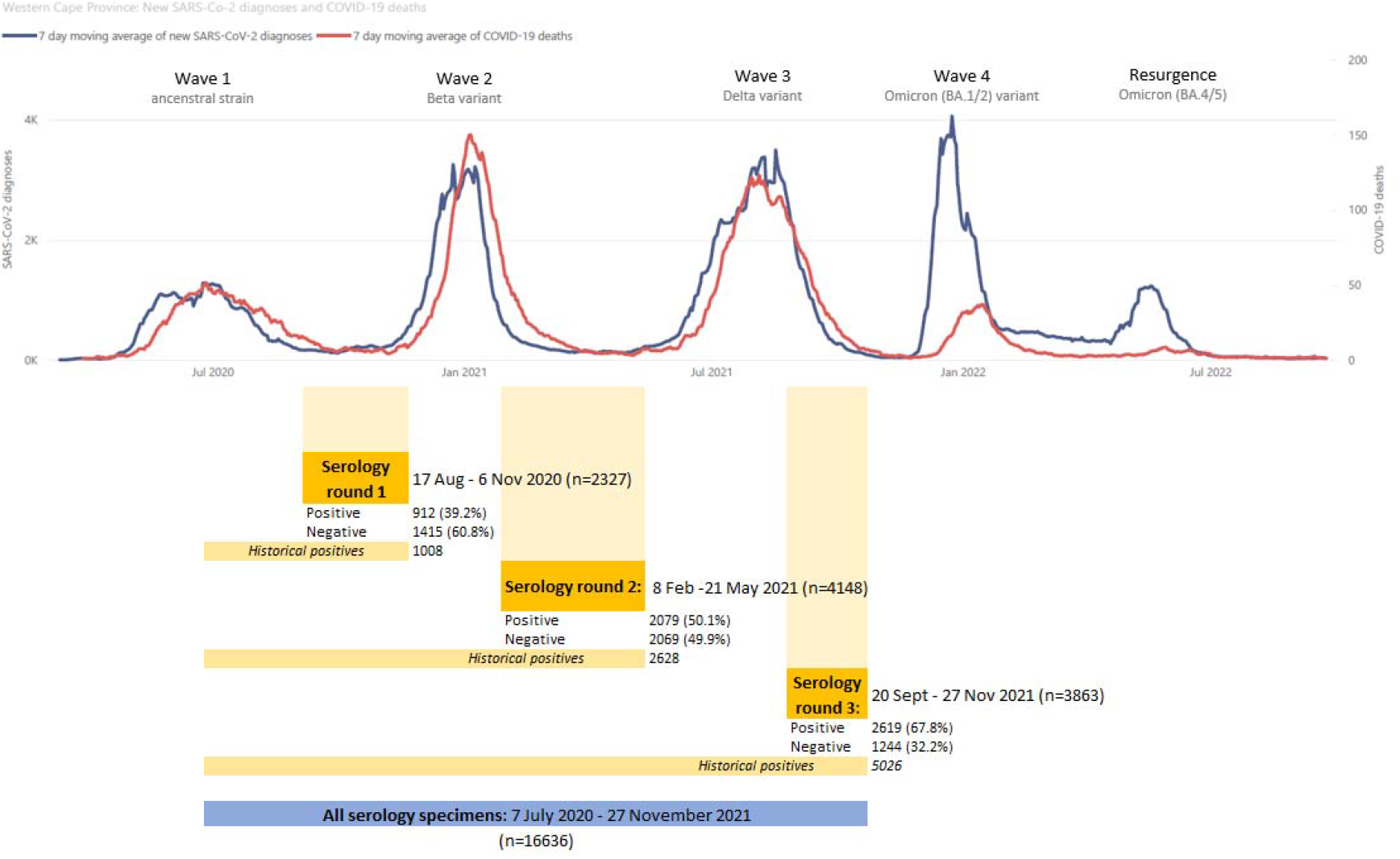
Line graph of new SARS-CoV-2 diagnoses and COVID-19 deaths in the Western Cape Province (March 2020 to August 2022), combined with the time periods of the included serology specimens to describe the relationship of the four COVID-19 waves relative to the serology testing rounds.

## Methods

### Source population

The city of Cape Town in South Africa had an estimated 4.6 million population in 2020 residing in eight subdistricts. Approximately 75% of the population does not have medical insurance and is dependent on public sector health services (13). The routine laboratory service of the public sector is provided by the National Health Laboratory Service (NHLS) laboratories.

We conducted three rounds of cross-sectional serosurveys in Cape Town. We selected consecutive convenience routine plasma samples collected for the purposes of: 1) antenatal blood grouping, 2) HIV viral load monitoring, 3) glycated haemoglobin testing and 4) paediatric in-patients and outpatients across various tiers of health facilities. Samples were retrieved from NHLS between April 2020 and November 2021. After the first round, antenatal sampling was dropped in favour of glycated haemoglobin samples to minimise sex and age biases.

### Assays

Prior infection with SARS-CoV-2 was assessed through detection of total anti-nucleocapsid (anti-N) antibody using the Elecsys™ anti-N SARS-CoV-2 assay (Roche, Geneva, Switzerland) on the Roche Cobas e601 platform according to manufacturer’s instructions. The assay is widely used for population seroprevalence studies and its performance was evaluated in South Africa where the specificity was 100% and sensitivity was 89% for detection of PCR-confirmed infection >30 days post diagnosis (14). The assay was deemed adequate for Section 21 use in serosurveys by the South African Health Products Regulatory Authority.

### PHDC and linkage of clinical data to serology results

The Western Cape Provincial Health Data Centre (PHDC) links patient administrative records, laboratory, pharmacy and vaccination data from routine electronic clinical information systems used in all public sector health facilities using a unique patient identifier (15). Since March 2020, the PHDC has collated information on all SARS-CoV-2 testing and related hospital admissions as part of routine government surveillance activities.

Serology results were linked at the PHDC to create an integrated dataset comprising a cohort of patients with all COVID-19 testing, vaccination status, admission and mortality data, patient demographics, subdistrict of last primary care visit, and known co-morbidities (HIV, tuberculosis history, diabetes, chronic kidney disease, hypertension, asthma/COPD). The data set was fully de-identified prior to being received for analysis.

Vaccination status was determined based on vaccination data obtained from the Electronic Vaccination Data System (EVDS). Vaccination status, which was defined as either single or double dose, was assessed at time of COVID-19 diagnosis, or at the peak of the wave if the individual did not test positive for COVID-19 during the wave of interest (3 January 2021 for second wave, 1 August 2021 for third wave and 19 December 2021 for fourth wave). Single dose vaccination could be with either Pfizer–BioNTech (BNT162b2) or Janssen/Johnson & Johnson (Ad26.COV2.S; J&J), while two dose vaccination was defined as being more than 14 days after receipt of any two doses of COVID-19 vaccines. Because of the timing and manner of vaccine roll out in South Africa, only two doses of Pfizer vaccinations were included in this latter group. Three or more doses was not assessed as these only became widely available in South Africa from March 2022, when the fourth wave was already ending (16), and so insufficient numbers of boosted individuals were available for this analysis.

### Descriptive and ecological analyses

Sero-positivity and 95% confidence estimates were calculated for the serology study population, stratified by age, sex, comorbidity and presumed subdistrict of residence based on location of last primary health care facility visit. Logistic regression to determine the odds ratio for having positive serology was also assessed, adjusted for age, sex, serology round and subdistrict of residence. To assess the impact of socio-economic status on SARS-CoV-2 infection and outcome, subdistrict seroprevalence estimates were correlated with subdistrict household income data and COVID-19 death rates. The proportion of low-income households in each of the subdistricts was determined using the latest available Census data from 2011 (17). Age-standardised COVID-19 death rates by subdistrict were determined using methodology which has previously been described (18).

### Individual linked analyses

For the linked analysis to estimate protection associated with prior infection and/or vaccination, three separate analyses for each wave were conducted. All specimens from serology preceding the wave of interest, as well as any additional historical positive specimens, were included in the analysis of that wave (Figure 1) - e.g. the first round of serology testing between 17 August and 6 Nov 2020, before the second wave, was used to look at the outcomes in the second wave. Prior infection was defined as individuals with positive anti-N serology in any period prior to the wave of interest, i.e., “historical” positive specimens were included in analyses of later waves, while only negative specimens from the serology round immediately preceding the wave could be included.

We categorised linked serology specimens according to their anti-N result and vaccination status. We used logistic regression to assess the outcomes of confirmed symptomatic infection (i.e., having a documented positive SARS-CoV-2 test in the Western Cape), being a non-severe case (i.e., a positive SARS-CoV-2 test but not admitted nor deceased), or a severe case (a positive SARS-CoV-2 test and admitted or deceased), adjusted for age, sex, subdistrict of residence and known comorbidities. We considered admissions within ±14 days of a positive antigen or PCR test as COVID-19 related and deaths up to 28 days after a positive COVID-19 test or within 14 days of discharge following a COVID-19 related admission to be COVID-19-related deaths.

### Study approval and funding

The study was approved by the University of Cape Town Human Research Ethics Committee (HREC REF 449/2020) and Stellenbosch University Human Research Ethics Committee (N20/08/051). Institutional approval was obtained from the National Health Laboratory Service and the Western Cape Department of Health. The study is funded by National Health Laboratory service, Western Cape Department of Health, Wellcome Trust, and in part by the Division of Intramural Research, NIAID, NIH. RJW is supported by the Francis Crick Institute which receives funding from Cancer Research UK (FC0010218), Medical Research Council (FC0010218), and Wellcome (FC0010218). He also received funding from Wellcome (203135,222754). For the purposes of open access the authors have applied a CC-BY public access copyright to any author-accepted manuscript arising from this submission. The funders had no role in the study design, collection and analysis of data and decision in submission for publication.

## Results

### Descriptive and ecological analyses results

Our study tested a total of 16 636 residual plasma samples across the first four waves of the SARS-CoV-2 epidemic in Cape Town. Of these, 10 338 specimens could be grouped into three rounds of serology testing in the inter-wave periods (Figure 1). Anti-N seroprevalence increased from 39.19% (95% confidence interval [95%CI] 37.23-41.19%) in the first round (i.e., after the first wave), to 50.12% (95%CI 48.60-51.64%) in the second round, and to 67.80% (66.31-69.25%) in the third round before the start of the fourth wave.

There was heterogeneity in seroprevalence across age, sex, comorbidities and subdistrict of residence (Figure 2). Higher seroprevalence was observed in individuals aged 17-49, women and people living with HIV compared to their older/younger, male and diabetic counterparts, respectively. These risk factors for seropositivity persisted after multivariable adjustment (Table 1). Compared to the first round of serology, the second and third rounds were associated with an adjusted odds ratio (aOR) for anti-N positivity of 1.96 (95%CI 1.76-2.19) and 3.89 (95%CI 3.47-4.36) respectively.

**Figure 2:**
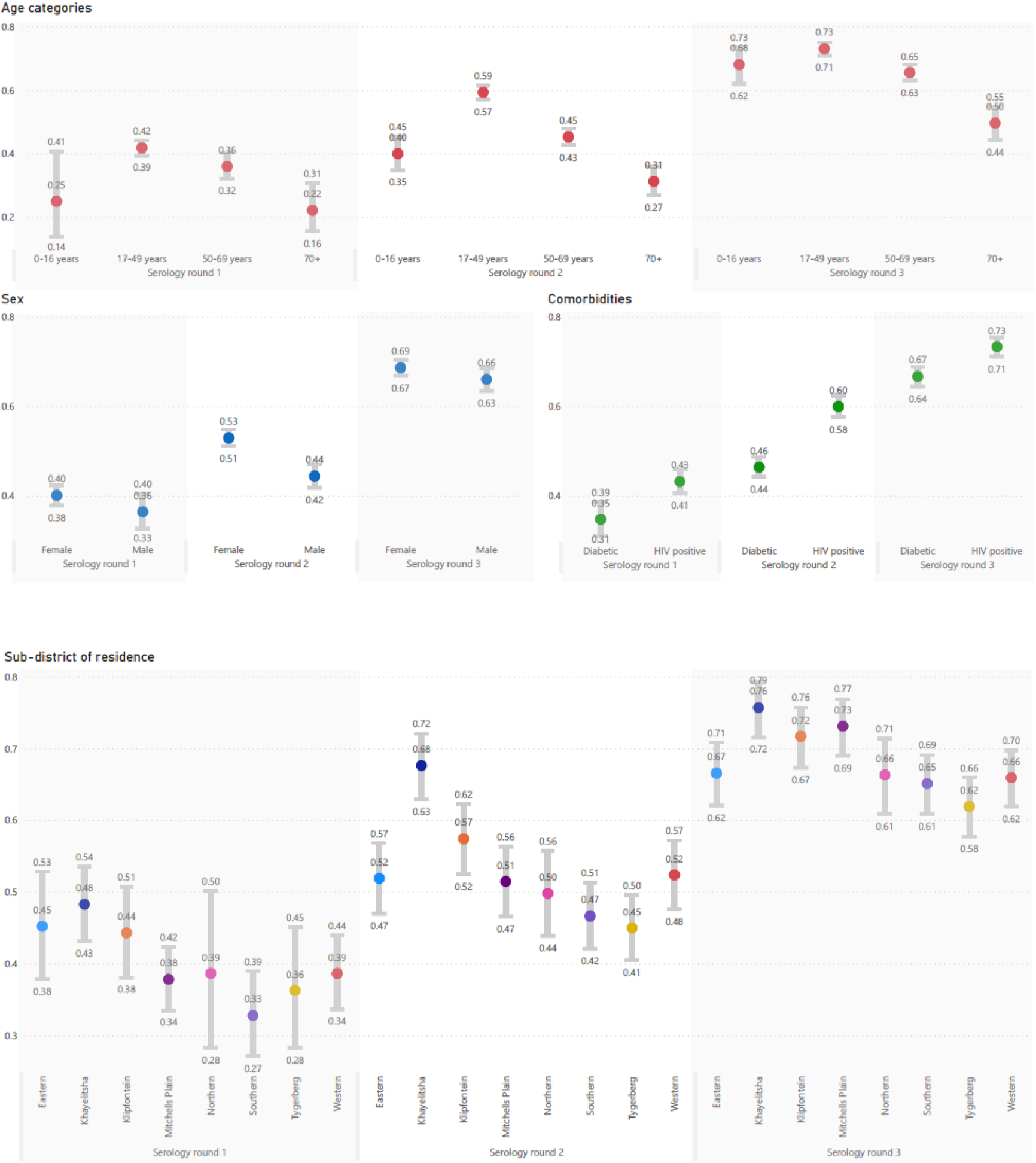
Anti-N seroprevalence and 95% confidence interval, grouped by serology round and age category, sex, comorbidity or subdistrict of residence.

**Table 1:** Logistic regression for the outcome of having anti-N positive serology.

|  |  | adjusted Odds<br>Ratio | (95% CI) |  |
| --- | --- | --- | --- | --- |
| Sex | Female | Ref |  |  |
|  | Male | 0.85 | 0.77 | 0.92 |
| Age category | 0-16 years | Ref |  |  |
|  | 17-49 years | 1.55 | 1.30 | 1.85 |
|  | 50-69 years | 1.04 | 0.87 | 1.24 |
|  | ≥ 70 years | 0.56 | 0.45 | 0.70 |
| Serology round | 1 | Ref |  |  |
|  | 2 | 1.96 | 1.76 | 2.19 |
|  | 3 | 3.89 | 3.47 | 4.36 |
| Subdistrict of<br>residence | Eastern | Ref |  |  |
|  | Khayelitsha | 1.59 | 1.33 | 1.90 |
|  | Klipfontein | 1.29 | 1.08 | 1.55 |
|  | Mitchells Plain | 1.08 | 0.91 | 1.28 |
|  | Northern | 0.93 | 0.75 | 1.14 |
|  | Southern | 0.90 | 0.75 | 1.07 |
|  | Tygerberg | 0.81 | 0.68 | 0.97 |
|  | Western | 0.99 | 0.84 | 1.18 |

While seroprevalence increased with time across Cape Town, there was a strong correlation between anti-N positivity and subdistrict poverty levels, with the poorest subdistrict, Khayelitsha, consistently having the highest seroprevalence (Figure 3a and 3b). This resulted in differing wave patterns in the different subdistricts (Figure 4). Northern subdistrict, the most affluent in the Cape Town Metro, had a relatively small first wave, but with increasingly large second and third waves. Khayelitsha, the poorest subdistrict, in contrast, had a very large first wave and then second and third waves of decreasing size. Figure 3c describes how low income subdistricts, which had the highest seroprevalence rates, also had the highest standardised COVID-19 death rates.

**Figure 3:**
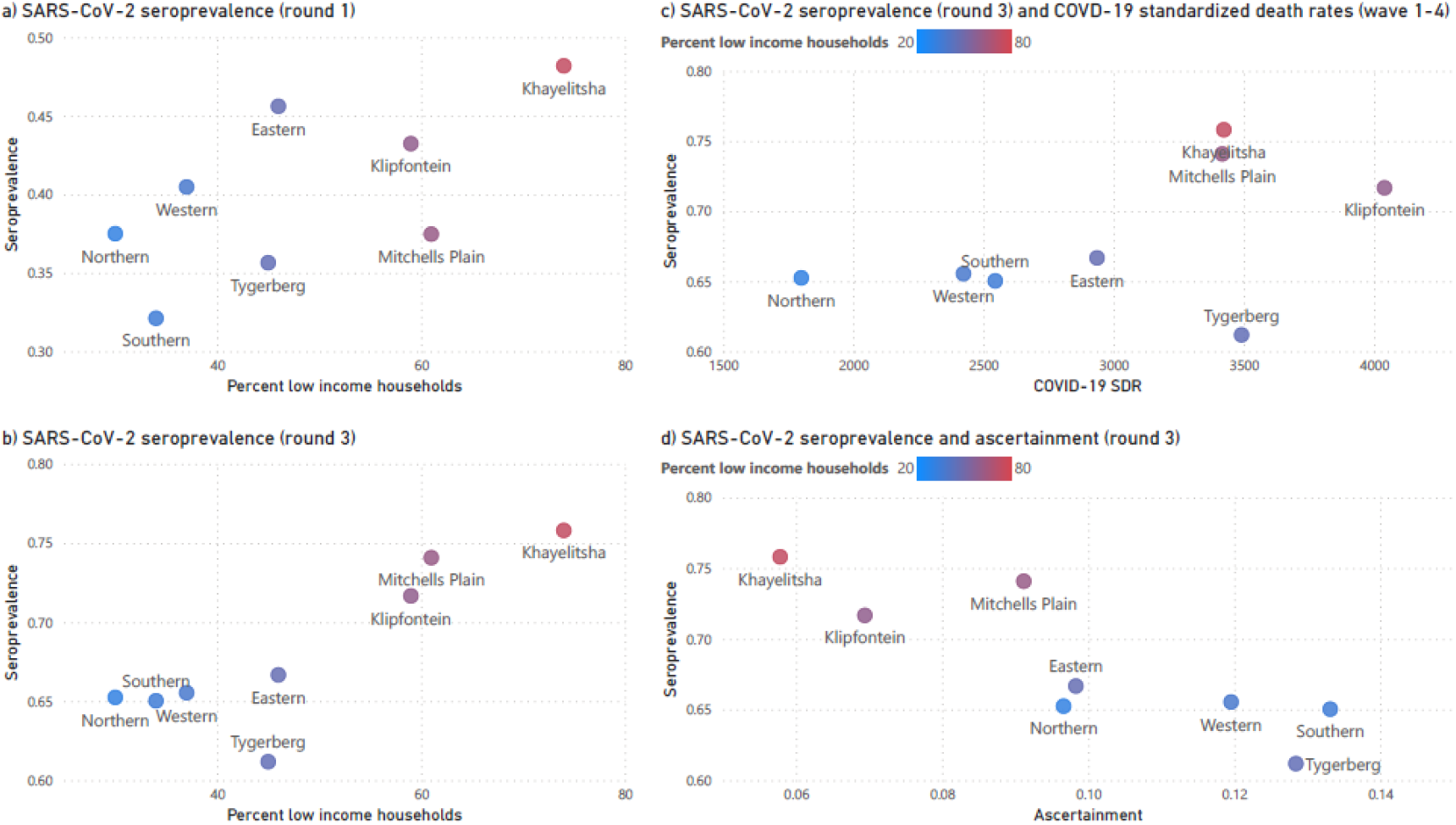
Anti-N seroprevalence scatter plots describing the association between (a) seroprevalence in serology round 1 (August – November 2020) with proportion of low-income households, (b) as above but for serology round 3 (September – November 2021), (c) seroprevalence in round 3 and age and sex-standardized COVID-19 deaths rates(SDR), combined for waves 1-4, and (d) seroprevalence in round 3 compared to the case ascertainment rate. In all the scatter plots, subdistrict is the unit of measurement, and is colour coded according to proportion of low-income households.

**Figure 4:**
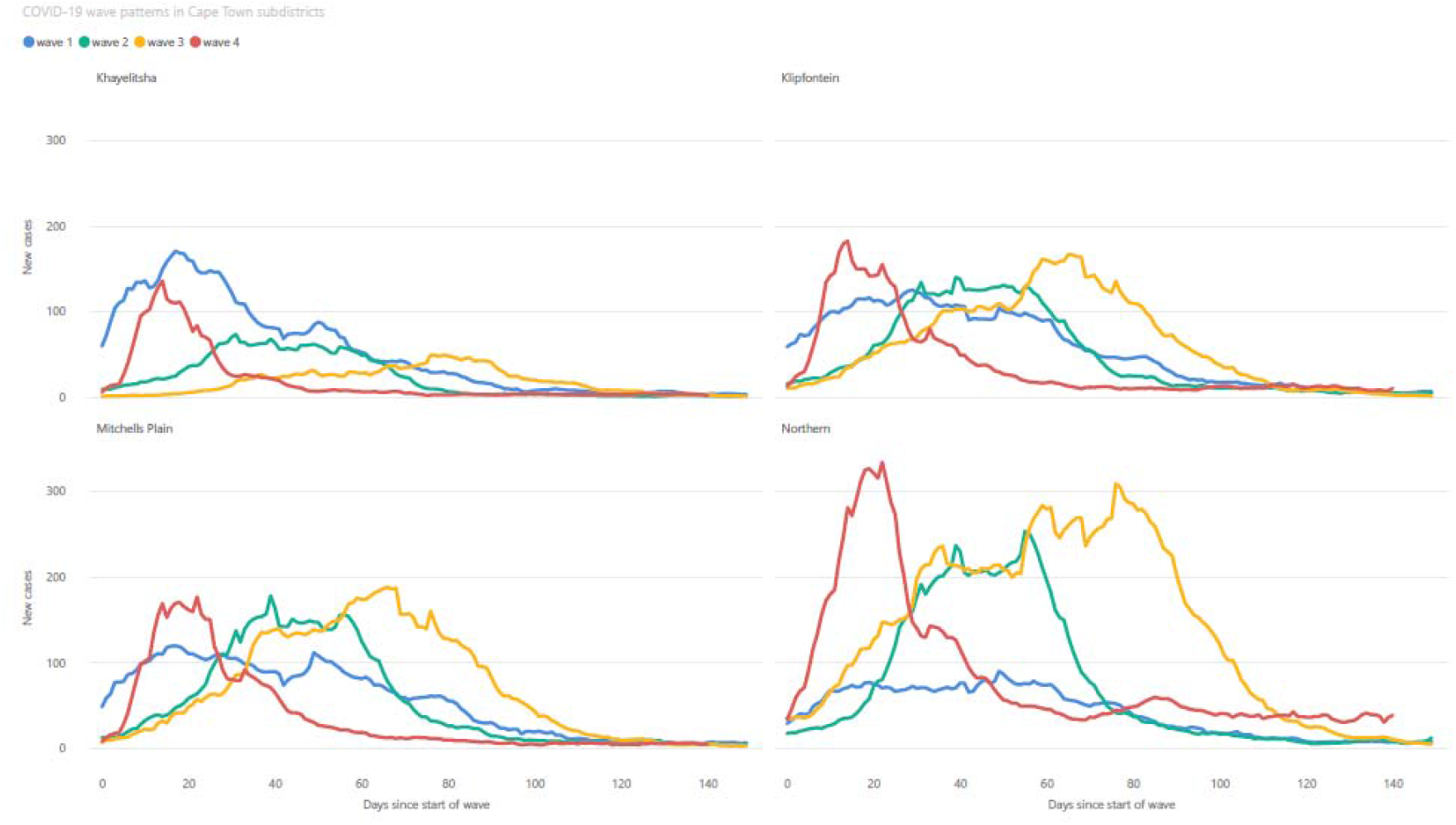
7 day moving average of new SARS-CoV-2 cases, in both the public and private sector, in four subdistricts of Cape Town (Northern, Khayelitsha, Klipfontein and Mitchells Plain), showing the differing patterns of COVID-19 waves. The wave patterns of the other four subdistricts, as well as Cape Town as a whole, are provided in the supplementary file.

The case ascertainment rate, that is the proportion of individuals with positive anti-N specimens who had a positive SARS-CoV-2 test recorded at any time before their positive serology, remained relatively stable over time (0.12, 0.12 and 0.10 in serology rounds 1,2 and 3 respectively – see Supplementary Table 1). Case ascertainment displayed marked variation by subdistrict, with the poorer subdistricts having lower ascertainment rates (Figure 3d). For example, in Khayelitsha, only 6% of cases were ascertained in both the first and third round of serology testing, compared to the Western subdistrict where 23% and 12% of cases were ascertained, respectively.

### Serology and linked subsequent infection/outcome results

For the linked analysis, there were 8 184 linked serology episodes available to assess outcomes in the fourth wave and 6 306 and 3 215 specimens for the third and second waves respectively. The distribution of antibody results and vaccination status, as well as outcomes, is shown in Table 2. As the vaccination programme in the general population only began in May 2021, everyone was unvaccinated in the second wave. In all waves, Anti-N antibody presence was associated with protection against subsequent confirmed symptomatic infection or having severe disease. Protection against severe disease was highest in the Delta wave (aOR of 0.02; 95%CI 0.00-0.11 for antibody positive vs negative in unvaccinated individuals), and lowest in the Omicron wave (aOR of 0.15; 95%CI 0.05-0.35). Vaccination (2 doses) in anti-N positive individuals further strengthened this protection against severe disease in the Omicron wave (aOR 0.07, 95%CI 0.01-0.35 for vaccinated anti-N positive vs unvaccinated anti-N negative).

**Table 2:**
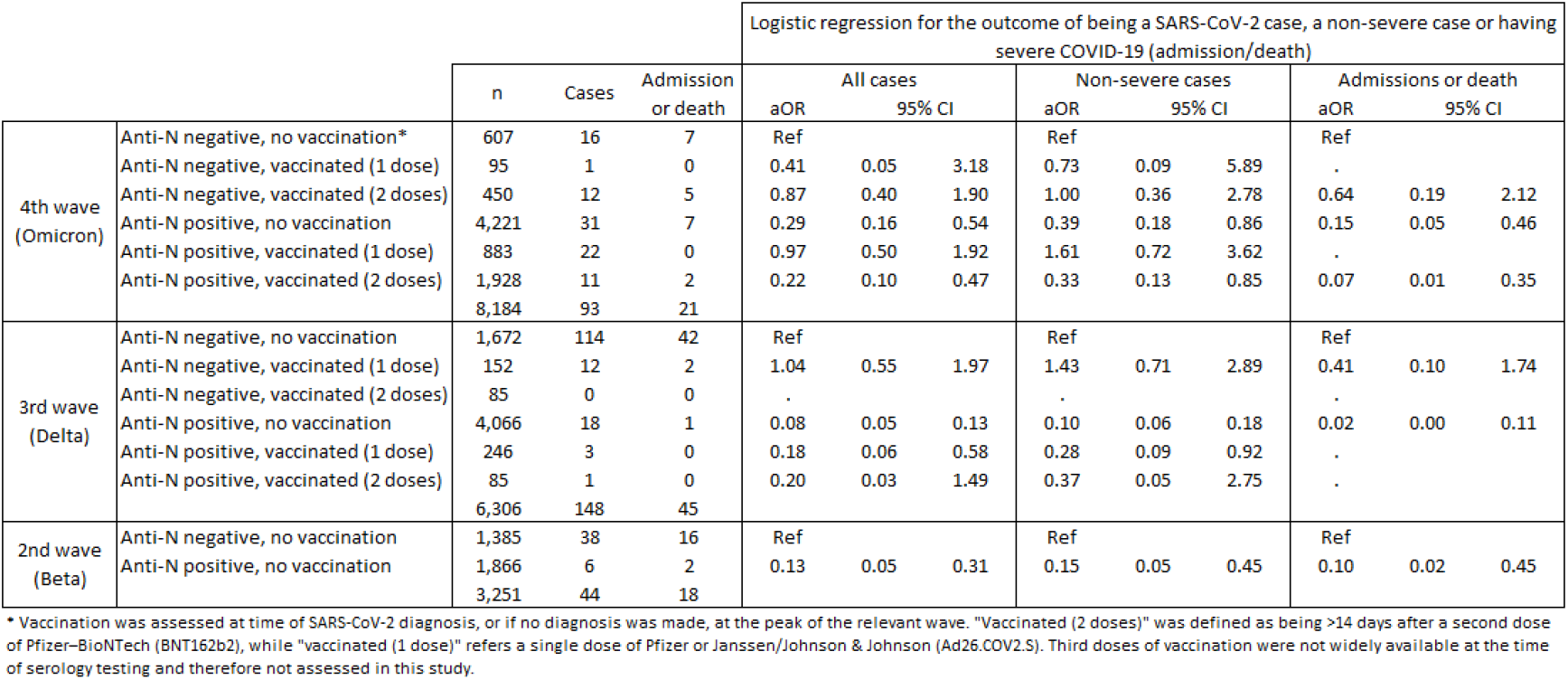
Linked serology analysis, showing the numbers of serology specimens and outcomes in each risk category, as well as logistic regression for the outcome of being a SARS-CoV-2 case, with and without severe disease. This analysis was adjusted for age category, sex, known comorbidities and subdistrict of residence.

## Discussion

Our seroprevalence data following three waves of the SARS-CoV-2 epidemic in Cape Town indicate that despite public health restrictions, the burden of infection in the community was high and anti-N seropositivity was strongly protective against severe disease during a subsequent infection, irrespective of variant.

Despite the differences in study design, our seroprevalence estimates were similar to other contemporaneous studies in South Africa (3–6) and Africa. Furthermore, we observed similar heterogeneity in the seroprevalence across demographic and social economic factors. HIV infection and less affluent subdistrict of residence, proxies for lower socio-economic status, were associated with higher seroprevalence across all three waves of infection. In a previous local study, these factors were also strongly associated with hospitalization and death, but at that point we could not differentiate the impact of high infection burden from other factors which might drive disease progression (19). The current study suggests that the higher mortality seen in poorer communities was substantially driven by higher risks of infection.

The epidemic wave asymmetry across subdistricts is best illustrated by Figure 4. The high levels of seropositivity from the first wave of COVID-19 infections in Khayelitsha likely provided protection from infection and severe disease in subsequent waves, despite the second and third waves being caused by variants of higher virulence (20). Omicron as an immune escape variant, however, resulted in a surge of infections in the fourth wave, even in areas like Khayelitsha.

Our study suggests that official case counts in Cape Town represented only ∼10% of all infections, and that the degree of under-ascertainment was highest in low-income communities. This case under-ascertainment has been observed in low- and middle-income countries and highlights the reality of unequal access to diagnosis and care. In addition, this under-ascertainment hampers the international comparison of epidemiology and pandemic trajectories, which in turn impacts resource allocation for future global response to emerging pathogens.

This study shows that anti-N positivity, a proxy for prior exposure, was associated with protection against severe disease, even in the context of an immune escape variant like Omicron, reducing the odds of hospital admission or death by 85%. The protection against confirmed symptomatic infection was more modest but the under-ascertainment of milder cases, particularly in the poorer communities, could have biased this estimate towards the null. This bias was further evident in the similar aORs for confirmed symptomatic infection and admission during the second wave. This was the period where testing capacity was severely constrained and there was greater bias towards testing individuals with risk factors for severe disease. Overall, our finding is broadly in line with studies included in a systematic review examining the issue of relative anti-N seropositivity protection (21).

While the fourth wave saw a sharp increase in SARS-CoV-2 cases, this did not result in a concomitant increase in related mortality (see line graph in Figure 1). Analyses from our setting have found that the Omicron variant was associated with less severe disease when compared to the Delta variant (20,21), but the low case ascertainment rates in this study complicates this interpretation (22).

Vaccination in addition to natural infection (anti-N seropositivity) further halved the risk of severe disease. A recent study also found that the combination of both vaccination and recent infection conferred the highest level of neutralizing antibodies against all variants, including Omicron sub-lineages (9). The broader, more potent immune response associated with this form of hybrid immunity likely plays an important role in the ongoing decoupling of COVID-19 cases and severe disease in South Africa.

While the linked analysis highlights how seropositivity to SARS-CoV-2 is protective for an individual in subsequent waves of COVID-19, and the ecological analyses demonstrates how it can be advantageous for communities to have high seroprevalence rates when they face subsequent waves of COVID-19, it is important to bear in mind that seropositivity is only achieved through the risk of severe illness and death during the first infection of SARS-CoV-2 (Figure 3c).

## Limitations

This study has several limitations. For the ecological analysis we were limited to using residual samples, of mostly chronically ill patients in care in the public sector, and it is uncertain how representative they are of the general population, with regards to exposure to infection and SARS-CoV-2 testing patterns. As suburb data was incomplete in the ecological analysis, we were restricted to using the subdistrict level, even though marked heterogeneity can exist within subdistricts.

In the linked serology analysis, misclassification of both the exposure and the outcome could have occurred, as the sensitivity of the antibody assay is imperfect, and with limited SARS-CoV-2 testing, particularly at the peaks of waves, there was under-ascertainment of cases. Related to testing, those who were vaccinated were more likely to access/present for diagnostic testing, resulting in health seeking behaviour being a confounder which may partly explain the lack a protective vaccination effect for the outcome of being a case.

While this study did find additional protection against severe disease from vaccination in those with prior infection, it is not a formal study of vaccine effectiveness. Most of the study population were diabetic or living with HIV, and the findings cannot directly be extrapolated to the general population. With such high background seroprevalence rates in the public sector population, the small numbers of individuals who remained anti-N negative in November 2021 might have a biological reason for not seroconverting, i.e., have suboptimal immune responses. This analysis also had low numbers of seronegative, vaccinated individuals prior to the fourth wave, and since anti-N positivity was associated with strong protection against severe disease, we were underpowered to adequately assess the additional benefit of vaccination amongst those who were already seropositive.

Another unavoidable limitation of this study is survivor bias, in that only those individuals who survive their first episode of COVID-19 can have their serology tested at a later time point and found to be seropositive.

## Conclusion

There is a complex interaction over time between socio-economic status and SARS-CoV-2 infection with differing variants, as well as the protection that both prior infection and vaccination confers, that has resulted in diverse COVID-19 wave patterns and experiences of the pandemic in different communities.

Seroprevalence to SARS-CoV-2 in Cape Town increased rapidly over three epidemic waves, with poorer communities having consistently higher prevalence rates. While seropositivity was highly protective in later infections at the individual level, such protection was only achieved at the cost of high initial population COVID-19 morbidity and mortality.

## Data Availability

All data produced in the present study are available upon reasonable request to the authors. The Western Cape Department of Health and Wellness evaluates research proposals for all research in the public health sector in the province, subject to standard research ethics, government approval and data governance prescripts. This includes those that draw on routine datasets like the current study.For more information

---

**Supplementary Table 1:** Case ascertainment rate for each round of serology testing, as well by subdistrict of residence, determined by calculating the proportion of positive anti-N antibody results, that had a laboratory confirmed SARS-CoV-2 diagnosis at any time prior to their serology result.

|  |  | Proportion<br>ascertainment | 95%CI |  |
| --- | --- | --- | --- | --- |
| Combined for serology<br>round 1 = 0.12 (95%CI<br>0.10-0.14) | Eastern | 0 | (no observations) |  |
|  | Khayelitsha | 0.06 | 0.04 | 0.11 |
|  | Klipfontein | 0.12 | 0.07 | 0.20 |
|  | Mitchells Plain | 0.12 | 0.08 | 0.18 |
|  | Northern | 0.07 | 0.02 | 0.24 |
|  | Southern | 0.22 | 0.14 | 0.32 |
|  | Tygerberg | 0.09 | 0.03 | 0.22 |
|  | Western | 0.23 | 0.16 | 0.30 |
| Combined for serology<br>round 2 = 0.12 (95%CI<br>0.11-0.14) | Eastern | 0.08 | 0.05 | 0.12 |
|  | Khayelitsha | 0.08 | 0.05 | 0.11 |
|  | Klipfontein | 0.10 | 0.07 | 0.14 |
|  | Mitchells Plain | 0.07 | 0.04 | 0.12 |
|  | Northern | 0.08 | 0.05 | 0.14 |
|  | Southern | 0.14 | 0.10 | 0.20 |
|  | Tygerberg | 0.12 | 0.08 | 0.17 |
|  | Western | 0.12 | 0.08 | 0.17 |
| Combined for serology<br>round 3 = 0.10 (95%CI<br>0.09-0.11) | Eastern | 0.10 | 0.07 | 0.14 |
|  | Khayelitsha | 0.06 | 0.04 | 0.09 |
|  | Klipfontein | 0.07 | 0.05 | 0.10 |
|  | Mitchells Plain | 0.09 | 0.07 | 0.13 |
|  | Northern | 0.10 | 0.06 | 0.15 |
|  | Southern | 0.13 | 0.10 | 0.17 |
|  | Tygerberg | 0.13 | 0.10 | 0.17 |
|  | Western | 0.12 | 0.09 | 0.16 |

**Supplementary Figure 1:**
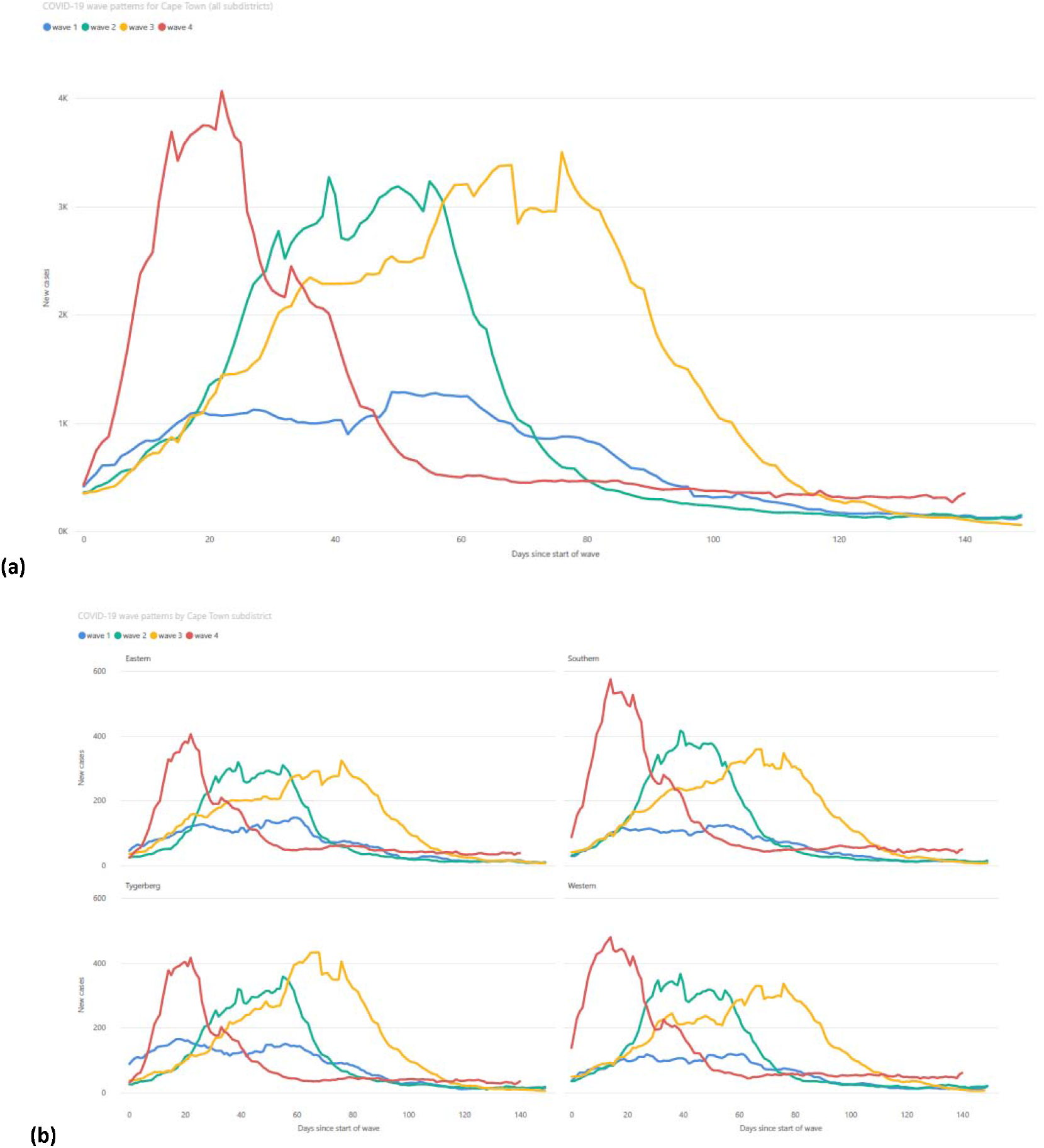
7 day moving average of new SARS-CoV-2 cases, in both the public and private sector, in the City of Cape Town as a whole (1a) and in the four remaining subdistricts of Cape Town (1b) not displayed in the main manuscript, showing the differing patterns of COVID-19 waves.

